# Low levels of the prognostic biomarker suPAR are predictive of mild outcome in patients with symptoms of COVID-19 - a prospective cohort study

**DOI:** 10.1101/2020.05.27.20114678

**Authors:** Jesper Eugen-Olsen, Izzet Altintas, Jens Tingleff, Marius Stauning, Hejdi Gamst-Jensen, Mette B Lindstrøm, Line Jee Hartmann Rasmussen, Klaus Tjelle Kristiansen, Christian Rasmussen, Jan O Nehlin, Thomas Kallemose, Ove Andersen

## Abstract

**OBJECTIVE:** To examine if baseline soluble urokinase plasminogen activator receptor (suPAR) can predict whether patients with COVID-19 symptoms will need mechanical ventilation during a 14-day follow-up. Furthermore, to examine differences in demographics, clinical signs, and biomarkers in patients tested either positive or negative for SARS-CoV-2.

**DESIGN:** Prospective cohort study including patients presenting with symptoms of COVID-19.

**SETTING:** Copenhagen University Hospital Amager and Hvidovre, Hvidovre, Denmark.

**PARTICIPANTS:** 407 patients presenting with symptoms of COVID-19 were included from the Emergency Department (ED). Patients were included from March 19 to April 3 and follow-up data was collected until April 17, 2020.

**MAIN OUTCOME MEASURES:** Primary outcomes were respiratory failure in patients presenting with symptoms of COVID-19 and in those with a positive SARS-CoV-2 RT-PCR test, respectively. Furthermore, we analysed differences between patients testing positive and negative for SARS-CoV-2, and disease severity outcomes in SARS-CoV-2 positive patients according to baseline suPAR.

**BACKGROUND:** Patients admitted to ED with clinical signs or symptoms of COVID-19 infection need a safe and quick triage, in order to determine if an in-hospital stay is necessary or if the patient can safely be isolated in their own home with relevant precautions. suPAR is a biomarker previously shown to be associated with adverse outcomes in acute medical patients. We aimed to examine if suPAR at baseline presentation is predictive of respiratory failure in patients presenting with symptoms of COVID-19. Furthermore, we examined demographic, clinical, and biochemical differences between SARS-CoV-2-positive and negative patients.

**RESULTS:** Among the 407 symptomatic patients, the median (interquartile range) age was 64 years (47-77), 58% were women, and median suPAR was 4.2 ng/ml (2.7-6.4). suPAR level below 4.75 ng/ml at admission ruled out respiratory failure during follow-up with an area under the curve (95% CI) of 0.89 (0.85-0.94) and a negative predictive value of 99.5%. Of the 407 symptomatic patients, 117 (28.8%) had a positive RT-PCR test for SARS-CoV-2 and presented with significant differences in vital signs, cell counts, and biomarkers compared to SARS-CoV-2 negative patients. In SARS-CoV-2 positive patients eligible for mechanical ventilation (N=87), 26 (30%) developed respiratory failure. Best baseline predictors of respiratory failure were suPAR with an area under the curve (95% CI) of 0.88 (0.80-0.95), EWS 0.84 (0.75-0.93), lactate dehydrogenase 0.82 (0.71-0.93), and C-reactive protein 0.80 (0.70-0.89).

**CONCLUSION:** SARS-CoV-2 affects several patient parameters underpinning the severe impact of the infection. A low suPAR level (<4.75 ng/ml) at baseline is a useful biomarker for aiding clinical decisions including discharge of patients presenting with symptoms of COVID-19.

## Introduction

The corona virus SARS-CoV-2 pandemic puts extraordinary pressure on hospital resources and capacity, especially for the Intensive Care Units (ICUs). Clinical and laboratory markers of potential respiratory failure are needed in order to safely and quickly distinguish between patients who will require admission to hospitals with ICU facilities, and patients who can be discharged to recover in another institution or in their private home.^1^

Several routine biomarkers have been shown to be associated with severe illness and mortality in patients with COVID-19. These include decreased white blood cell count, lymphocyte and platelet counts, and increased alanine aminotransferase (ALAT), creatinine kinase, lactate dehydrogenase (LDH), ferritin, C-reactive protein (CRP), interleukin-6, D-dimer^2^ and soluble urokinase plasminogen activator receptor (suPAR)^3^. suPAR is a prognostic biomarker that has been measured in acutely admitted medical patients at the Emergency Department (ED) at Copenhagen University Hospital Hvidovre since 2013. ^4,5^

Early and safe discharge of patients with symptoms of COVID-19 can lower the use of hospital resources, such as protective equipment, free up bed capacity, and lower the pressure on hospital staff. The main complication of patients with COVID-19 is the sudden development of respiratory failure, and safe discharge would require little or no risk of development of respiratory failure.

There are several reasons why suPAR may be a suitable marker for discharge or admission of patients with symptoms of COVID-19: First, suPAR is a marker of chronic inflammation, and patients with high suPAR have a chronically activated immune system that is less capable of fighting disease. Second, suPAR is elevated in elderly with comorbidities and identifies the frail patients. In unselected acute medical patients, suPAR is a strong marker of readmission and mortality.^2 4 5^ Third, suPAR has been investigated in several viral infections, including HIV, hepatitis C, Crimean-Congo hemorrhagic fever, and hantavirus. In all cases, suPAR is associated with clinical severity and mortality.^6-9^ Fourth, suPAR is elevated and associated with disease severity in patients with respiratory diseases, including pneumonia^10^ and chronic obstructive pulmonary disease,^11^ and predicts risk of acute respiratory distress syndrome in patients with sepsis.

We aimed to investigate whether suPAR and other biomarkers and clinical symptoms can aid in identifying patients with low risk of respiratory failure when presenting with symptoms of COVID-19. We aimed to identify a clinical cut-off value for suPAR for patients with symptoms of COVID-19 (Obs-COVID-19) and for those tested positive in reverse transcription – polymerase chain reaction (RT-PCR) testing for SARS-CoV-2. Furthermore, we aimed to determine whether the existing cut-off of 6 ng/ml is associated with disease severity in those who tested negative for SARS-CoV-2. Finally, we compared biomarkers and clinical signs between SARS-CoV-2-positive and negative Obs-COVID-19 patients.

## Methods

A single center prospective cohort study at Copenhagen University Hospital Hvidovre. Patients with mild symptoms and without need for clinical evaluation for admission were tested for SARS-CoV-2 in a temporary COVID-19 clinic. Only patients with more severe symptoms were referred to the ED and had blood drawn and were included in the study. Patient inclusion took place in the period from March 19^th^ until April 3^rd^, and with follow-up through April 17^th^, 2020.

### Laboratory measurements

#### Corona virus testing

SARS-CoV-2 testing was carried out on material obtained from expectorate, nasopharyngeal suction, tracheal secretion, lumbar puncture, BAL, or graft from pharynx using a RealStar® SARS-CoV-2 RT-PCR Kit RUO (Altona Diagnostics, Hamburg, Germany) adapted to a Roche flow system. The limit of detection was reported to be 50 copies of RNA.

#### Biomarker measurements

Blood samples were obtained at admission (within first 24 hours). White blood cell counts, haemoglobin, creatinine, CRP, ALAT, LDH, and bilirubin were measured using a COBAS 8000 analyzer (Roche Diagnostics, Mannheim, Germany). Cell counts (leukocyte, thrombocyte, lymphocyte, and neutrophils) were measured using flow cytometry on a Sysmex XN 9000 (Sysmex Corporation, Kobe, Japan).

Plasma suPAR was measured using suPARnostic Quick Triage point-of care test (ViroGates, Birkerød, Denmark) and quantified using an aLF reader (QIAGEN, Hilden, Germany). Blood for this test (EDTA, 4 ml) was drawn on arrival and centrifuged for 3 minutes. The test provides a result in 20 minutes.

suPAR was measured real time 24/7, but the result was not available to the attending physicians as it was unknown whether suPAR added value in triage of patients with symptoms of COVID-19. Data were transferred from electronic patient health records into a Research Electronic Data Capture program (REDCap), and quality control of data was performed by three medical doctors (IA, MS, and JT).^4, 14^.

### Clinical signs at admission

At admission, clinical signs and Early Warning Score (EWS; based on systolic blood pressure, pulse, respiratory rate, saturation, and body temperature) were assessed.

Furthermore, at the time of admission, patients were asked for number of days with symptoms of infection before presenting at the ED. Symptoms included sore throat, cough (productive, non-productive), body pain, fatigue, headache, dizziness, nausea/vomit, fever, abdominal pain, obstipation, diarrhoea, dysuria, dyspnea, chest pain, arthralgia, cramps, chills, and hemoptysis. Baseline comorbidities were recorded and include chronic obstructive pulmonary disease, asthma, type 1 diabetes, type 2 diabetes, hypertension, heart failure, diagnosed coronary disease, cancer – active, cancer - non-active, chronic renal failure, chronic liver disease, other lung disease, other heart disease, other chronic infectious disease, other inflammatory disease, alcohol abuse, or other relevant disease.

### Incident organ dysfunction or worsening of chronic organ dysfunction during hospitalization

Organ dysfunction during hospitalization was defined as either kidney failure (defined as dialysis initiated), liver failure (defined as INR >1.6 for patients without anticoagulant therapy, and/or ALAT >400 U/L, and/or bilirubin >50 umol/L, and/or albumin less than 15 g/L); lung failure (defined as > 10 L oxygen per minute and respiratory rate ≥28/minute or need of mechanical ventilation); heart failure (defined as Troponin T above 100 ng/ml, echo with newly decreased pump function or lowering of pump function, or acute heart failure that is not caused because of hypoxia following interruption of ventilation).

### Patient and Public Involvement

Our study did not involve patient or public involvement as there were no impact/change in clinical practice during the study as we did not know whether the clinical signs or biomarker measurements could impact on the outcome of the patients with symptoms of COVID-19.

### Data sharing statement

Upon reasonable request, and following regulatory approval, data may be shared.

### Outcomes

#### Primary outcome

Respiratory failure, defined as endotracheal intubation and mechanical ventilation in patients eligible for intensive care treatment. One patient developed sudden RF and died before transfer to the ICU and was included in respiratory failure analysis.

#### Secondary outcomes

All outcomes evaluated at 14 days follow-up. Discharged within 24 hours, still admitted, died, need of oxygen, Continuous Positive Airway Pressure (CPAP), ventilator, Extra Corporal Membrane Oxygenation (ECMO), vasopressor drugs, developed organ failure, kidney failure, liver failure, lung failure, heart failure.

### Statistical analysis

A schematic presentation of the analysis is shown in supplementary table 1.

Continuous and categorical variables are presented as median (interquartile range; IQR) and n (%), respectively.

Comparisons of baseline parameters between patients eligible/not eligible for intubation and SARS-COV-2 status (positive/negative) were done by chi-squared test, fishers exact test, or Wilcoxon sum rank test.

Optimal cut-off points for suPAR, age and all other biomarker measures in relation to respiratory failure was calculated by separate receiver operating characteristics (ROC) analysis. The Youden index (maximization of sensitivity and specificity) was used to determine optimal cut-off values. Area under the curve (AUC) with 95 % confidence interval (CI), sensitivity, specificity, positive predictive value (PPV) and negative predictive value (NPV) for cut-off values were reported for all ROC analysis. Comparisons of outcome parameters between suPAR below/above cut-off from ROC analysis were done by chi-squared test or fishers exact test.

Because of multiple testing when comparing baseline or outcome variables Bonferroni correction was used, scaling the significance level according to the number of tests performed. Since different numbers of variables were tested in the comparisons the significance level also varied, and specific significance levels is listed in the tables.

Incidence plots for time to development of respiratory failure in all patients with symptoms of COVID-19 and eligible to receive intensive care treatment and for those with additionally a positive test for SARS-CoV-2 within suPAR groups were made, using cut-off values from the ROC analysis to group patients. The statistical program R v3.60 (R Foundation for Statistical Computing, Vienna, Austria) was used for analysis and figures.

### Ethics

The database and collection of clinical data and entering into REDCap was approved by the Danish Data Protection Agency (record no. P-2020-513) and by the Danish Patient Safety Authority (record no. 31-1521-319).

## Results

### Baseline characteristics of patients presenting with symptoms of COVID-19

A total of 407 patients presenting at the ED with symptoms of COVID-19 (mainly cough and fever) from March 19^th^ to April 3^rd^, 2020, were included in the study. Median age was 64 years (IQR: 47-77), and 172 (42,3%) were women. Median suPAR level at admission was 4.1 ng/ml (IQR: 2.7-6.2). At baseline, clinicians deemed whether patients were eligible to be intubated, if necessary, during follow-up. Out of the 407 Obs-COVID-19 patients, 327 were assessed eligible to receive intensive care treatment if needed, while 80 were not due to frailty and/or the patient requested not to receive mechanical ventilation. Table 1 shows baseline clinical and biochemical characteristics of all Obs-COVID-19 patients and characteristics of patients eligible for intubation or not.

**Table 1.** Baseline data for all patients, patients eligible for intubation, and patients not eligible for intubation. All data were collected at first presentation in the ED. P value refers to differences between patients eligible for intubation or not. Values are presented as median (interquartile range (IQR)) or N (%). suPAR: soluble urokinase plasminogen activator receptor. CRP: C-reactive protein. Bonferroni-corrected significance level = 0.0019

| Variable | Obs-COVID-19 patients | Eligible patients | Non-eligible patients | P value |
| --- | --- | --- | --- | --- |
| N | 407 | 327 | 80 |  |
| Age (years) | 64 (47 - 77) | 60 (43 - 73) | 81 (73 - 86) | <0.001 |
| Female (%) | 235 (57.7%) | 186 (56.9%) | 49 (61.3%) | 0.56 |
| Male (%) | 172 (42.3%) | 141 (43.1%) | 31 (38.8%) |  |
| Number of comorbidities | 2 (1 - 3) | 1 (1 - 3) | 3 (2 - 4) | <0.001 |
| Systolic blood pressure | 135 (123 - 150) | 136 (124 - 150) | 133 (121 - 150) | 0.63 |
| Diastolic blood pressure | 81 (72 - 89) | 82 (74 - 90) | 77 (67 - 87) | 0.0059 |
| Pulse (bpm) | 86 (76 - 98) | 86 (76 - 97) | 84 (76 - 100) | 0.82 |
| Respiratory rate (bpm) | 20 (18 - 24) | 20 (17 - 22) | 22 (20 - 28) | <0.001 |
| Saturation (%) | 97 (95 - 99) | 98 (96 - 100) | 95 (93 - 97) | <0.001 |
| Temperature (°C) | 37.5 (36.9 - 38.1) | 37.4 (36.9 - 38) | 37.6 (37.1 - 38.3) | 0.068 |
| Early Warning Score (EWS) | 2 (0 - 4) | 2 (0 - 3) | 4 (1 - 7) | <0.001 |
| suPAR (ng/ml) | 4.1 (2.7 - 6.2) | 3.8 (2.6 - 5.7) | 5.8 (4.3 - 7.8) | <0.001 |
| Leukocyte count (10<sup>9</sup>/L) | 8.1 (6.1 - 10.9) | 8 (6.3 - 10.5) | 9.8 (5.4 - 12.1) | 0.21 |
| Thrombocyte count (10<sup>9</sup>/L) | 248 (192 - 302) | 250 (194 - 301) | 238 (177 - 307) | 0.52 |
| Lymphocyte count (10<sup>9</sup>/L) | 1.5 (0.9 - 2.2) | 1.5 (1.0 - 2.3) | 1.1 (0.7 - 1.6) | <0.001 |
| Neutrophil count (10<sup>9</sup>/L) | 5.3 (3.6 - 8.0) | 5.3 (3.7 - 7.6) | 7.1 (3.4 - 9.5) | 0.020 |
| Haemoglobin | 8.4 (7.6 - 9.1) | 8.5 (7.8 - 9.2) | 7.9 (6.8 - 8.8) | <0.001 |
| CRP (ug/ml) | 17.5 (2.6 - 74.5) | 11.0 (1.8 - 63.5) | 43.5 (14.2 - 120) | <0.001 |
| Lactate dehydrogenase (U/L) | 203 (177 - 265) | 198 (175 - 259) | 230 (193 - 277) | 0.0045 |
| Alanine aminotransferase | 23 (17 - 35) | 23 (17 - 35) | 23 (16 - 35) | 0.77 |
| (U/L) |  |  |  |  |
| Creatinine (μmol/l) | 77 (62 - 96) | 74 (62 - 92) | 92 (74 - 117) | <0.001 |
| Bilirubin (μmol/l) | 7 (5 - 11) | 7 (5 - 11) | 8 (5 - 11.25) | 0.124 |
| Active smoker | 84 (21.3%) | 73 (23.2%) | 11 (13.8%) | 0.00426 |
| Ex-smoker | 150 (38%) | 107 (34%) | 43 (53.8%) |  |
| Never smoked | 161 (40.8%) | 135 (42.9%) | 26 (32.5%) |  |
| Symptoms before admission<br>(days) |  |  |  |  |
| 0-1 days | 75 (19%) | 57 (17.7%) | 18 (25%) | 0.71 |
| 2-3 days | 85 (21.6%) | 68 (21.1%) | 17 (23.6%) |  |
| 4-5 days | 49 (12.4%) | 39 (12.1%) | 10 (13.9%) |  |
| 6-7 days | 51 (12.9%) | 44 (13.7%) | 7 (9.7%) |  |
| 8-10 days | 30 (7.6%) | 24 (7.5%) | 6 (8.3%) |  |
| 11-13 days | 17 (4.3%) | 14 (4.3%) | 3 (4.2%) |  |
| 14-15 days | 40 (10.2%) | 35 (10.9%) | 5 (6.9%) |  |
| 16+ days | 47 (11.9%) | 41 (12.7%) | 6 (8.3%) |  |
| Cough no | 144 (35.4%) | 111 (33.9%) | 33 (41.2%) | 0.274 |
| Cough yes | 263 (64.6%) | 216 (66.1%) | 47 (58.8%) |  |
| Fever no | 200 (49.1%) | 159 (48.6%) | 41 (51.2%) | 0.767 |
| Fever Yes | 207 (50.9%) | 168 (51.4%) | 39 (48.8%) |  |

### suPAR and risk of respiratory failure in Obs-COVID-19 patients eligible for intubation

During follow-up, 26 patients of the 327 eligible Obs-COVID-19 patients ended up receiving intubation and mechanical ventilation. In ROC curve analysis, the AUC for baseline suPAR for predicting respiratory failure was 0.89 (95% CI: 0.85-0.94). The Youden index for suPAR was 4.75 ng/ml. This provided a high NPV of 99.5%, a PPV of 23%, specificity of 71%, and sensitivity of 96%. A cumulative incidence plot of risk of respiratory failure in eligible patients with symptoms of COVID-19 with baseline suPAR above or below 4.75 ng/ml is shown in Fig. 1.

**Fig 1.**
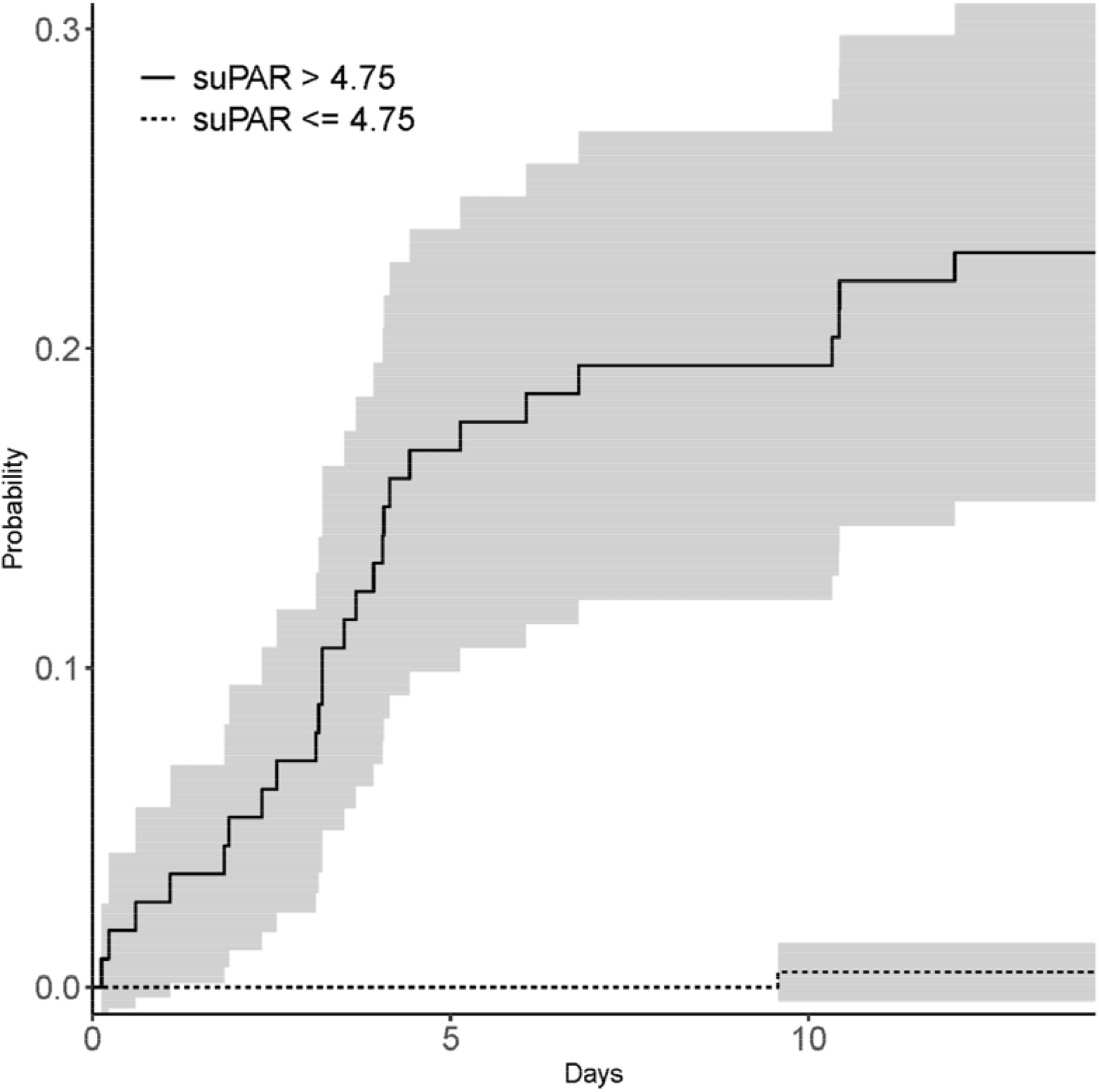
Incidence plot of eligible patients presenting with symptoms of COVID-19 and developing respiratory failure during 14 days of follow-up. Patients were divided into groups with baseline suPAR below or above 4.75 ng/ml. Grey areas indicate 95% confidence intervals.

### Baseline differences between SARS-CoV-2 positive and negative patients

Of the 407 Obs-COVID-19 patients, 117 (28.8%) tested positive for SARS-CoV-2 in the RT-PCR assay. Of these 117 with confirmed SARS-CoV-2, 22 had tested positive before arrival to the ED, 92 were tested positive in their first test in the ED, and 3 needed additional testing to confirm infection. The 117 patients who tested positive for SARS-CoV-2 differed on several clinical and biochemical parameters from the 290 patients who tested negative. Baseline characteristics of vital signs, symptoms, white blood cell counts, and biomarkers stratified by patients who tested positive or negative for SARS-CoV-2 are shown in table 2. Patients with a positive SARS-CoV-2 test were older, had lower blood saturation and higher EWS, higher suPAR and CRP levels, and lower leukocyte, thrombocyte, lymphocyte, and neutrophil counts (all p<0.001). There were fewer active smokers among patients who tested positive for SARS-CoV-2 (7.1% versus 27.0%, p<0.001).

**Table 2:** Baseline clinical and biochemical markers according to SARS-CoV-2 testing result. Parentheses refer to median (IQR) or N (%). Bonferroni-corrected significance level = 0.0019

| Variable | SARS-CoV-2 negative | SARS-CoV-2 positive | P value |
| --- | --- | --- | --- |
| N | 290 | 117 |  |
| Age (years) | 61 (43 - 77) | 71 (58 - 80) | <0.001 |
| Female (%) | 167 (57.6%) | 68 (58.1%) | 1 |
| Male (%) | 123 (42.4%) | 49 (41.9%) |  |
| Number of comorbidities | 2 (1 - 3) | 2 (1 - 3) | 0.29 |
| Systolic blood pressure | 136 (124 - 152) | 133 (122 - 143) | 0.107 |
| Diastolic blood pressure | 82 (74 - 90.75) | 79 (69 - 87) | 0.0206 |
| Pulse (bpm) | 86 (75 - 98) | 86 (77 - 96) | 0.971 |
| Respiratory rate (bpm) | 20 (17.25 - 23) | 20 (18 - 24) | 0.035 |
| Saturation (%) | 98 (96 - 100) | 96 (94 - 98) | <0.001 |
| Temperature (Celsius) | 37.3 (36.8 - 37.7) | 38 (37.3 - 38.6) | <0.001 |
| Early Warning Score | 2 (0 - 4) | 3 (1 - 5) | <0.001 |
| suPAR (ng/ml) | 3.7 (2.6 - 5.5) | 5.3 (4 - 7.7) | <0.001 |
| Leukocyte count (E9/L) | 9.1 (7.0 - 11.7) | 6.1 (4.5 - 7.6) | <0.001 |
| Thrombocyte count (E9/L) | 260 (208 - 311) | 199 (163 - 265) | <0.001 |
| Lymphocyte count (E9/L) | 1.7 (1.1 - 2.5) | 1 (0.72 - 1.4) | <0.001 |
| Neutrophil count (E9/L) | 6 (4.1 - 8.75) | 4.2 (3 - 5.9) | <0.001 |
| hemoglobin | 8.4 (7.6 - 9.2) | 8.3 (7.5 - 9.1) | 0.526 |
| CRP (ug/ml) | 8.4 (1.6 - 48.5) | 44 (18 - 99) | <0.001 |
| Lactate dehydrogenase (U/L) | 192 (173 - 229) | 269 (201 - 359) | <0.001 |
| Alanine aminotransferase (U/L) | 21.5 (16 - 31) | 28.5 (21 - 45.3) | <0.001 |
| Creatinine (mmol/l) | 74 (62 - 93) | 84 (67 - 102) | 0.00694 |
| Bilirubin (mmol/l) | 7 (5 - 12) | 7 (5 - 10) | 0.861 |
| Active smoker | 76 (27%) | 8 (7.1%) | <0.001 |
| Ex-smoker | 104 (36.9%) | 46 (40.7%) |  |
| Never smoked | 102 (36.2%) | 59 (52.2%) |  |
| Number of days with symptoms before admission | 68 (24.2%) | 7 (6.2%) | <0.001 |
| 2-3 days | 59 (21%) | 26 (23%) |  |
| 4-5 days | 31 (11%) | 18 (15.9%) |  |
| 6-7 days | 36 (12.8%) | 15 (13.3%) |  |
| 8-10 days | 15 (5.3%) | 15 (13.3%) |  |
| 11-13 days | 6 (2.1%) | 11 (9.7%) |  |
| 14-15 days | 28 (10%) | 12 (10.6%) |  |
| 16+ days | 38 (13.5%) | 9 (8%) |  |
| Cough -No | 109 (37.6%) | 35 (29.9%) | 0.177 |
| Cough -Yes | 181 (62.4%) | 82 (70.1%) |  |
| Fever - No | 164 (56.6%) | 36 (30.8%) | <0.001 |
| Fever - Yes | 126 (43.4%) | 81 (69.2%) |  |

### Baseline clinical and biochemical parameters and their association to development of respiratory failure in SARS-CoV-2 positive patients

A total of 87 SARS-CoV-2 positive patients (74%) were deemed eligible to receive mechanical ventilation if needed whereas 28 were not. During follow-up, 26 (30%) of the 87 developed respiratory failure (25 were intubated and 1 died before intubation). Of the 25 intubated patients, 11 (44%) died during the 14-day follow-up. Best baseline predictors of respiratory failure were suPAR with an AUC (95% CI) of 0.88 (0.80-0.95), EWS 0.84 (0.75-0.93), LDH 0.82 (0.71-0.93), and CRP 0.80 (0.70-0.89). The Youden index for suPAR was 5.2 ng/ml (table 3). Other baseline parameters including age, cell counts, and other routine biomarkers had an AUC of 0.65 or lower (table 3). ROC curves for the four best predictive biomarkers are shown in fig. 2.

**Table 3:** Baseline data were used to calculate ROC AUCs for respiratory failure (ventilation) for patients eligible for mechanical ventilation (N=87).

| **Mechanical ventilation during follow-up\*** | Number of patients with data available N (%) | COVID-19 test positives AUC (95% CI) | Cut-off (Youden index) | Specificity | Sensitivity | PPV | NPV |
| --- | --- | --- | --- | --- | --- | --- | --- |
| Age | 87 (100) | 0.57 (0.44-0.71) | 66 | 0.61 | 0.69 | 0.43 | 0.82 |
| EWS score | 87 (100) | 0.84 (0.75-0.93) | 3.5 | 0.77 | 0.77 | 0.59 | 0.89 |
| suPAR (ng/ml) | 87 (100) | 0.88 (0.80-0.95) | 5.2 | 0.7 | 0.88 | 0.56 | 0.93 |
| Leukocytes (10<sup>9</sup>/L) | 87 (100) | 0.59 (0.45-0.74) | 7.2 | 0.77 | 0.54 | 0.5 | 0.8 |
| Thrombocytes (10<sup>9</sup>/L) | 87 (100) | 0.62 (0.49-0.74) | 191 | 0.49 | 0.77 | 0.39 | 0.83 |
| Lymphocytes (10<sup>9</sup>/L) | 87 (100) | 0.65 (0.52-0.79) | 0.63 | 0.89 | 0.46 | 0.63 | 0.79 |
| Neutrophil (10<sup>9</sup>/L) | 87 (100) | 0.63 (0.49-0.77) | 5.8 | 0.82 | 0.5 | 0.54 | 0.79 |
| Haemoglobin (mmol/l) | 87 (100) | 0.53 (0.38-0.67) | 9.1 | 0.8 | 0.46 | 0.5 | 0.78 |
| CRP (mg/L) | 86 (99) | 0.80 (0.70-0.89) | 64 | 0.67 | 0.8 | 0.5 | 0.89 |
| Lactate dehydrogenase (U/L) | 72 (83) | 0.82 (0.71-0.93) | 310 | 0.74 | 0.84 | 0.53 | 0.93 |
| Alanine aminotransferase (U/L) | 86 (99) | 0.60 (0.45-0.74) | 50 | 0.84 | 0.44 | 0.52 | 0.78 |
| Creatinine (μmol/l) | 85 (98) | 0.62 (0.48-0.76) | 94 | 0.76 | 0.54 | 0.5 | 0.79 |
| Bilirubin (μmol/l) | 86 (99) | 0.50 (0.37-0.63) | 3.5 | 0.13 | 1 | 0.32 | 1 |

ROC: receiver operator curve, AUC: Area under the curve, CI: Confidence Interval, PPV: positive predictive value, NPV: negative predictive value. suPAR: soluble urokinase plasminogen activator receptor, EWS: Early warning score, CRP: C-reactive protein

**Fig 2.**
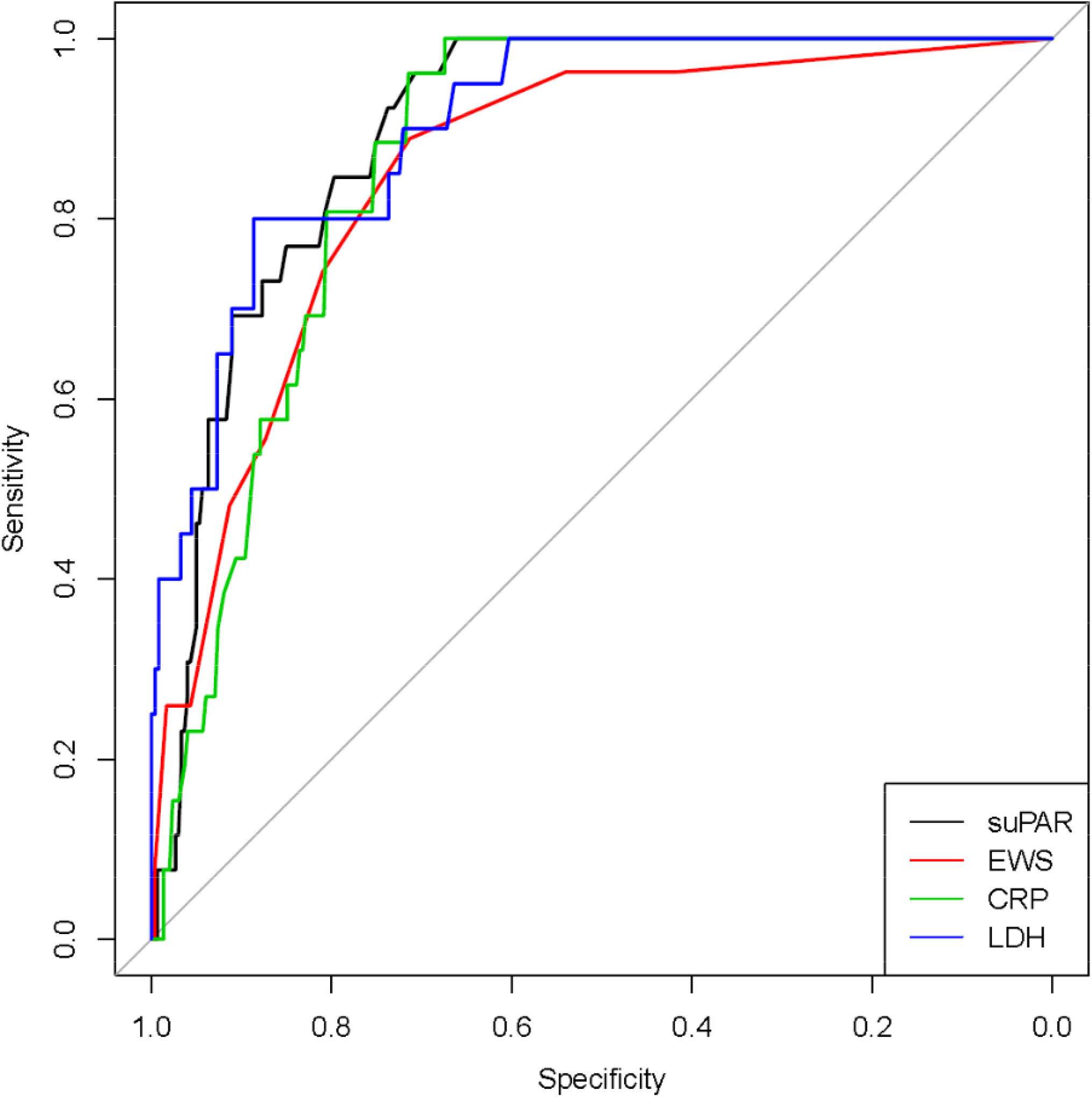
Biomarkers at baseline as predictors of respiratory failure in eligible patients (n=87) with a positive test for SARS-CoV-2 during 14 days of follow-up. suPAR: soluble urokinase plasminogen activator receptor, EWS: Early Warning Score, CRP: C-reactive protein, LDH: Lactate dehydrogenase. AUCs are shown in table 3.

### Elevated baseline suPAR levels in SARS-CoV-2 positive patients are associated with severe outcomes

The Youden analysis found that a suPAR level of 5.2 ng/ml was the optimal cut-off for development of respiratory failure in the eligible SARS-CoV-2 positive patients. A cumulative incidence plot stratified according to baseline suPAR level above or below 5.2 ng/ml and development of respiratory failure is shown in Figure 3. To determine whether suPAR is associated with outcomes in addition to respiratory failure, such as acute kidney injury, we divided patients according to baseline suPAR below or above 5.2 ng/ml in table 4. Patients with suPAR above 5.2 ng/ml were more severely affected by the infection, with more patients still being admitted at day 14 (46.3% versus 8.7%), and were more often receiving pressor drugs (48.8% versus 4,3%) and more required dialysis (26.8% versus 0%)(all p<0.001, table 4).

**Table 4:**
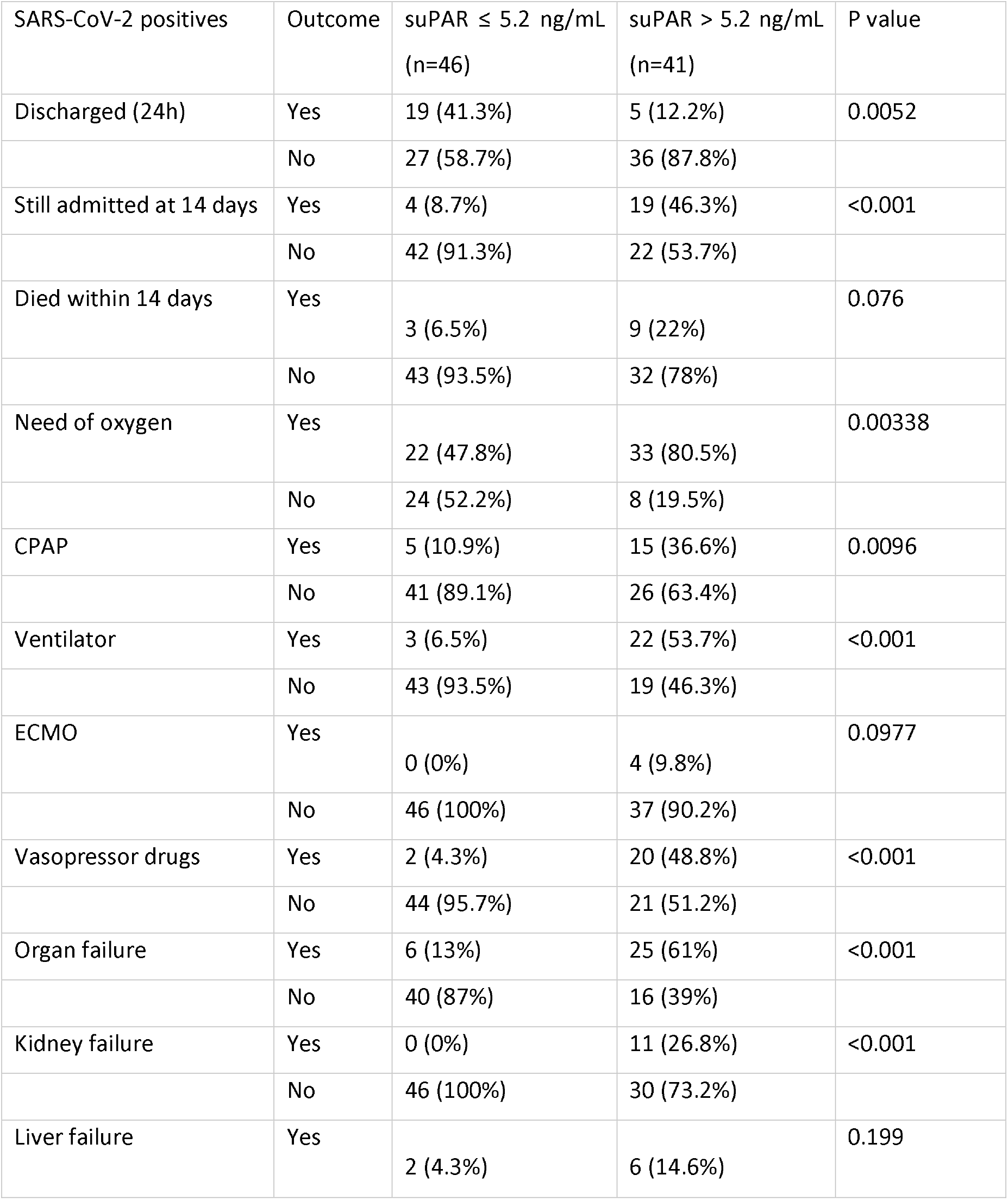
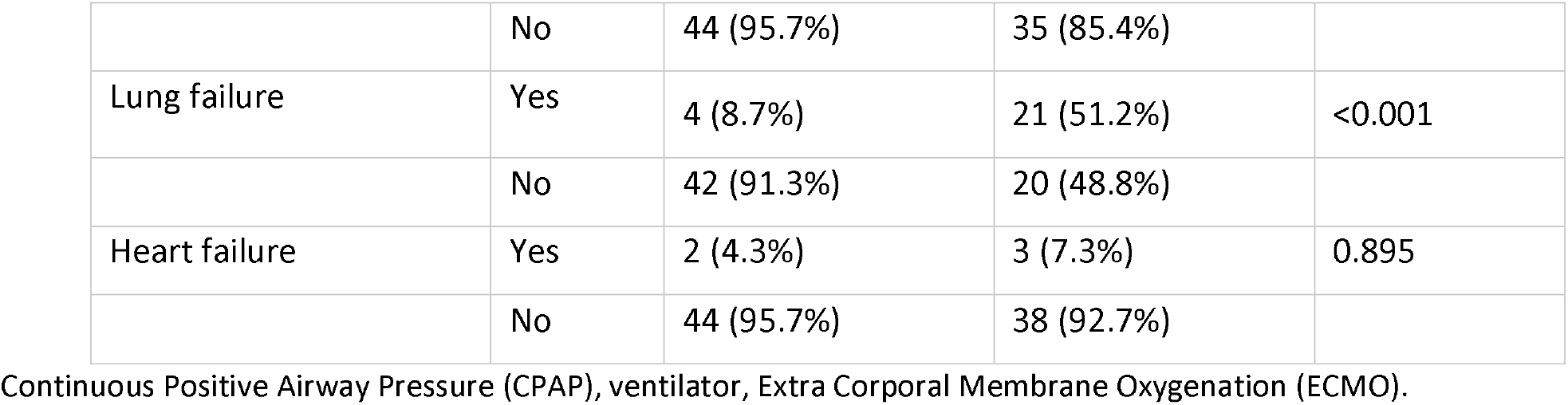
Outcome among patients tested positive for SARS-CoV-2 and stratified according to baseline suPAR below or above 5.2 ng/ml (cut-off calculated as Youden index). Bonferroni-corrected significance level = 0.0036.

**Fig 3.**
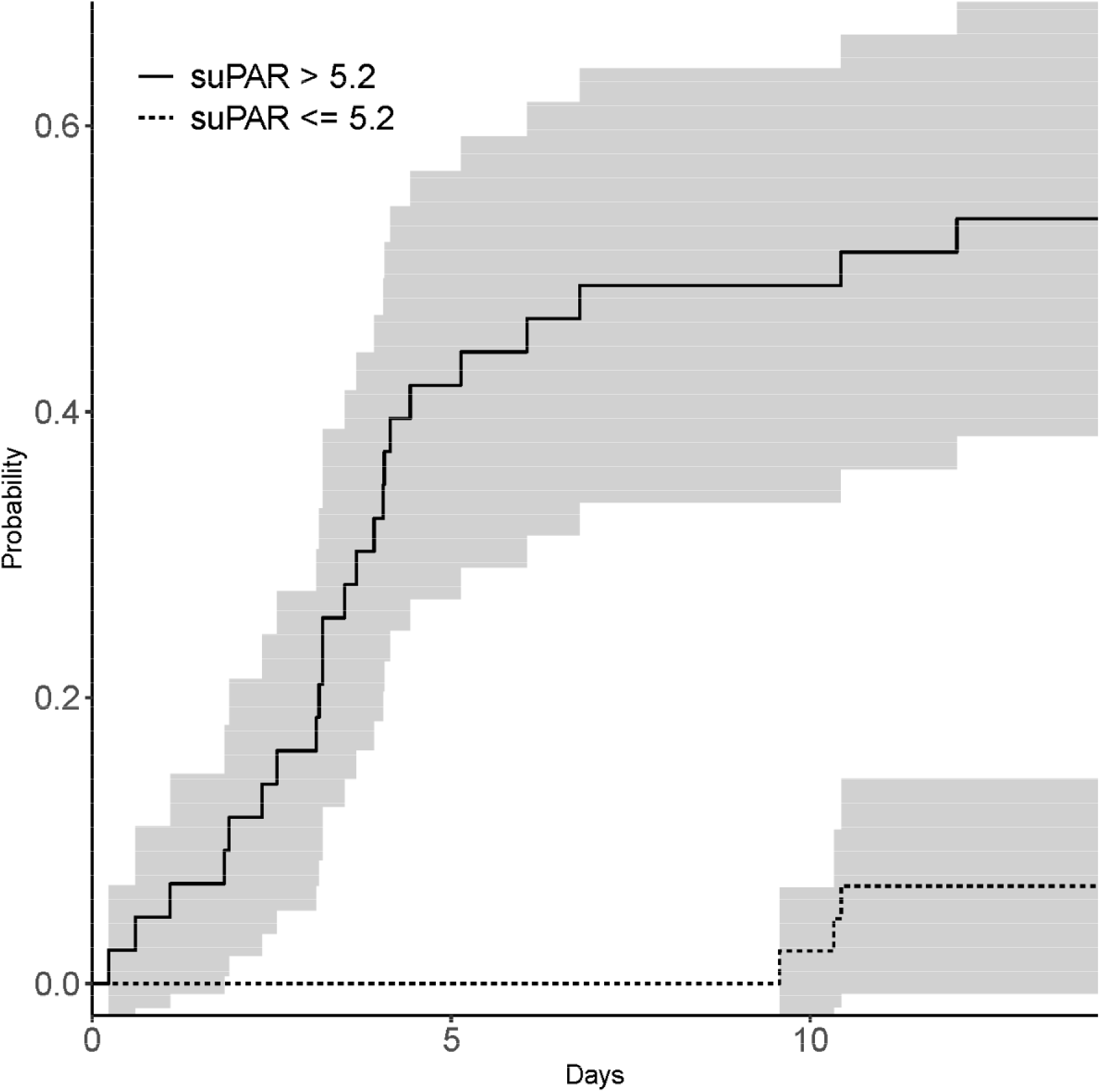
Incidence plot of eligible patients presenting with positive test for SARS-CoV-2 and developing respiratory failure (N=26) during 14-days of follow-up. Patients were divided into groups with baseline suPAR below or above 5.2 ng/ml (Youden index for SARS-CoV-2 positive patients). Grey areas indicate 95% confidence intervals.

## Discussion

In the ED at Hvidovre Hospital, approximately 1/3 of the patients presenting with symptoms of COVID-19 were tested positive, and 1/3 of these developed respiratory failure requiring intubation and mechanical ventilation. It should be emphasized that only patients with more severe symptoms are referred to the ED for blood works and examination. We have previously shown that low suPAR can is associated with low risk of adverse outcomes for acute medical patients. ^4,5^. Our present results suggest that suPAR may also be useful for risk stratification of patients with symptoms of COVID-19, as suPAR below 4.75 ng/ml had a negative predictive value of 99.5% of requiring ventilator support during the 14 days of follow-up. Obs-COVID-19 patients with low suPAR might therefore be discharged to facilities without ventilator access.

### Major differences between Obs-COVID-19 patients who tested positive or negative for SARS-CoV-2

We observed several differences with respect to clinical signs, biomarkers and cell counts between patients who tested positive and negative for SARS-CoV-2, respectively. Those who tested positive for SARS-CoV-2 were older, had lower blood saturation, and strikingly lower cell counts as well as elevated suPAR, CRP, and LDH levels, underpinning the severe impact of this viral infection on multiple parameters. These clinical and biochemical differences are worth noting as the SARS-CoV-2 testing may not always be immediately available, and testing may produce false negatives.^15^ Fewer SARS-CoV-2 positives were active smokers, which has also been observed in a French study,^16^ but we did not investigate whether this was due to older age or other parameters. Further studies are needed to determine whether active smoking or social habits related to smoking protects against SARS-CoV-2 infection.

### Eligibility for intubation

At baseline, clinicians evaluated whether a patient was eligible for intubation if necessary. Deciding which patients are eligible for intubation is more relevant than ever. SARS-CoV-2 infection causes severe lung injury with patients often being intubated for weeks, often on their stomach and with high oxygen levels and high pressure on the ventilator that may impact on cardio function. Furthermore, a high patient load may also necessitate selection of patients that have the best chance of surviving mechanical ventilation. We provide the clinical and biochemical characteristics according to patient eligibility in our hospital. Those not eligible for ICU treatment were, in general, elderly fragile comorbid patients.

### Elevated suPAR is associated with increased risk of respiratory failure in SARS-CoV-2 positive patients

In patients who tested positive for SARS-CoV-2, approximately 1/3 developed respiratory failure and were intubated and ventilated during the 14 days follow-up. Among the biomarkers examined in this study, we identified baseline suPAR as a good baseline predictor of respiratory failure during follow-up (AUC of 0.88), suggesting that suPAR can aid in the identification of COVID-19 patients at low or high risk of respiratory failure. Our study is in accordance with the findings of a recent Greek/American study on COVID-19 patients that found a 16-fold increased risk of intubation in patients with baseline suPAR above 6 ng/ml.^3^

### Baseline suPAR as a marker of disease progression and disease severity in acute medical patients

Since 2013, our hospital has measured suPAR in acute medical patients. We have shown that elevated suPAR is a strong marker of disease severity, readmission, and mortality.^4 5^ However, as an unspecific biomarker reflecting overall patient disease severity, low suPAR seems particular useful as a discharge marker. A randomized controlled study found more patients were discharged when clinicians had access to suPAR.^17^ The present study find that elevated suPAR is also associated with disease severity and outcomes in patients presenting with symptoms of COVID-19, as shown by the higher number of long-term hospital admission, increased number of patients developing respiratory and organ failures.

A recent position paper from the Hellenic Sepsis study group suggests suPAR below 4 ng/ml for discharge and above 6 ng/ml for further examination.^18^ While suPAR above 6 ng/ml may be a good cut-off level indicating need for clinical attention and further examination, suPAR below 6 ng/ml may be too high for safe discharge. The Youden Index identified a suPAR cut-off of 4.75 ng/ml for respiratory failure in Obs-COVID-19 patients in the current study, in agreement with safe discharge after thorough clinical examination if the patient has a suPAR level below 4 ng/ml.

### Perspectives of the findings

Currently, many potential treatments and drugs that may aid COVID-19 patients are in the pipeline or being tested. We propose that trials include patients with high risk of negative outcomes as the possible benefit may outweigh the potential side effects in low risk patients. The first trial only including patients with elevated suPAR has been initiated (suPAR-guided Anakinra Treatment for Validation of the Risk and Management of Respiratory Failure by COVID-19, ClinicalTrials.gov Identifier: NCT04357366).

We show that patients who tested positive for SARS-CoV-2 had elevated suPAR, and a suPAR level of 5.2 ng/ml had the maximum sensitivity and specificity for respiratory failure. COVID-19 patients with suPAR above 5.2 ng/ml also had increased risk of developing acute kidney injury. Recent studies have shown that elevated suPAR is not only associated with, but may also be causative in the development of acute kidney injury.^19 20^ This raises the question of whether anti-suPAR antibody treatment may lower the overzealous immune activation observed in severe COVID-19 patients and protect against AKI.^19^ Further studies are needed to examine this.

Another question raised by our findings is whether the suPAR level is a risk factor for COVID-19 susceptibility? First, suPAR is a marker of chronic inflammation, and patients and even individuals without disease are prone to disease development and disease progression with elevated suPAR.^19 21^

Second, a chronically activated immune system may also allow for higher risk of infection with virus, as virus may have an easier time evading the immune system, settling, and replicating. Third, suPAR is the soluble form of the urokinase receptor, uPAR, a receptor that promotes plasminogen activation to plasmin. Plasmin proteolytically breaks down excess fibrin to generate D-dimer, a biomarker also observed to be associated with outcome in COVID-19. Plasmin may cleave the S protein of SARS-CoV-2, which increases its infectivity and virulence.^22^ Thus, an elevated suPAR before infection may increase susceptibility to SARS-CoV-2 infection, but further studies are needed to verify this.

## Limitations

This is a single center study and larger studies with longer follow-up are needed to confirm suPAR as a predictive marker in COVID-19. Another limitation is that we were not able to compare suPAR to D-dimer, IL-6 and ferritin which have also been shown to be associated with respiratory failure in COVID-19 patients.^2, 23^

In conclusion, suPAR is a marker of disease development in patients presenting at the ED with symptoms of COVID-19. The SARS-CoV-2 virus affects several biomarkers of the infected patients. Patients with elevated baseline suPAR were at increased risk of respiratory failure during 14-day follow-up. Patients with symptoms of COVID-19 and a suPAR level less than 4-.75 ng/ml can be discharged following thorough clinical examination to facilities without ventilator access.

## Data Availability

Upon reasonable request, and following regulatory approval, data may be shared.

## Acknowledgements

We wish to thank all the patients involved in this study. We thank Marianne Falck for excellent technical assistance and Linda Camilla Andresen for effective coordination and organization.

## Authors’ contribution

JEO, IA, JT, and OA has contributed to the conception of the study. IA, MS, HGJ, and JT have performed data collection. MBL and TK has performed data management. IA and TK have carried out the analyses and takes responsibility for the integrity and accuracy of the data analysis. All authors contributed with interpretation of data, critically revision of the manuscript, and read and approved the final version. The corresponding author attests that all listed authors meet authorship criteria and that no others meeting the criteria have been omitted. JEO is the guarantor of the study.

## Ethical approval

The study was approved by the Danish Data Protection Agency (record no. P-2020-513) and the Danish Patient Safety Authority (record no: 31-1521-319). There was no need of approval from the National Committee on Health Research Ethics since only registries were used.

## Consent for publication

Not applicable

## Availability of data and material

The data that support the findings of this study are available from REDCap database, but restrictions apply to the availability of these data, which were used under license for the current study, and therefore they are not publicly available

## Transparency declaration

The lead authors (the manuscript’s guarantor) affirms that the manuscript is an honest, accurate, and transparent account of the study being reported; that no important aspects of the study have been omitted; and that any discrepancies from the study as planned (and, if relevant, registered) have been explained.

## Dissemination to participants and related patient and public communities

No study participants were involved in the preparation of this article.

**Supplementary figure 1.**
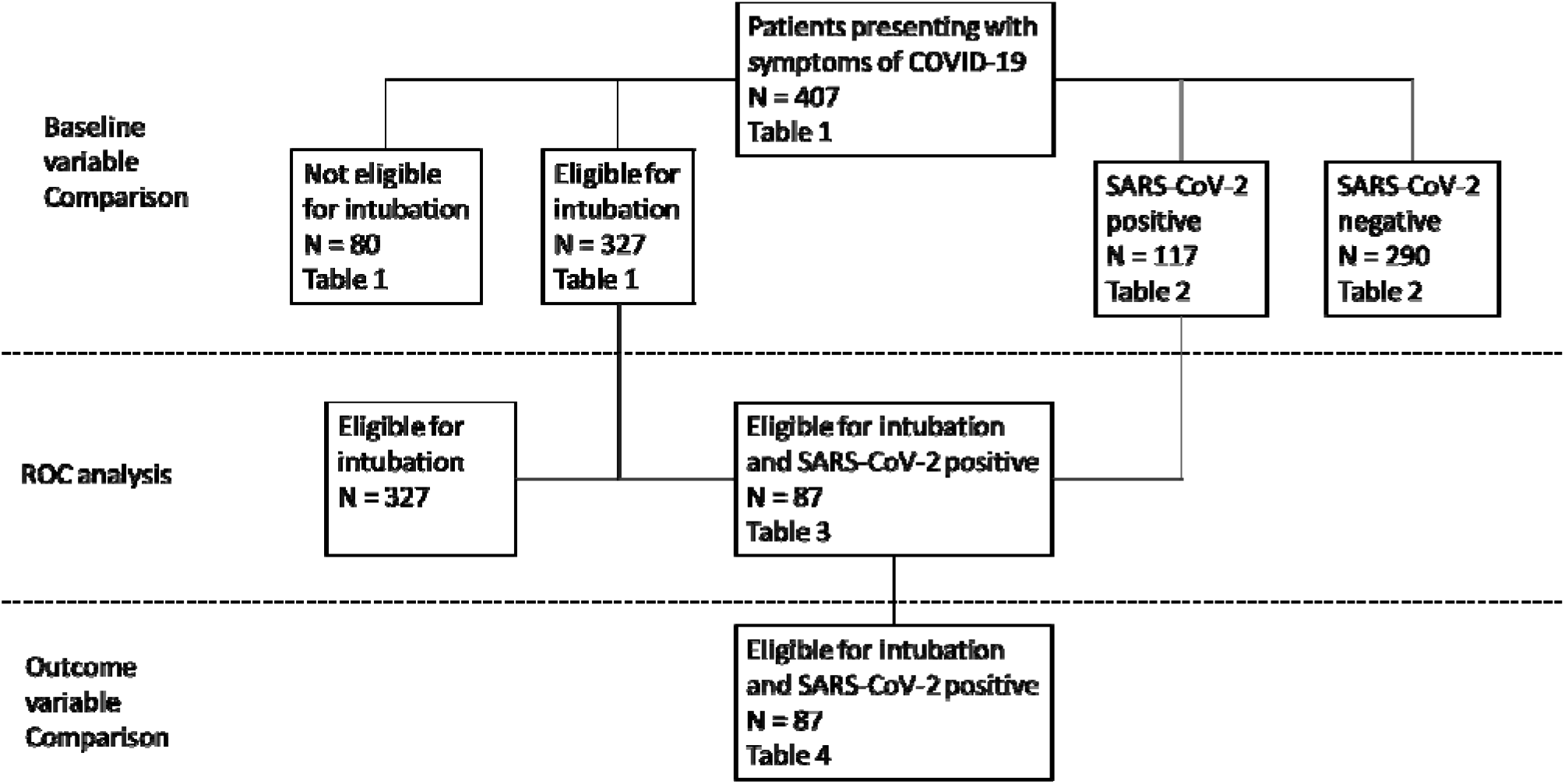
Schematic presentation of analyses carried out.

## References

1. Vitacca M, Nava S, Santus P, et al. Early consensus management for non-ICU ARF SARS-CoV-2 emergency in Italy: from ward to trenches. Eur Respir J 2020 doi: 10.1183/13993003.00632-2020 [

2. Henry BM, de Oliveira MHS, Benoit S, et al. Hematologic, biochemical and immune biomarker abnormalities associated with severe illness and mortality in coronavirus disease 2019 (COVID-19): a meta-analysis. Clin Chem Lab Med 2020 doi-10.1515/cclm-2020-0369

3. Rovina N, Akinosoglou K, Eugen-Olsen J, et al. Soluble urokinase plasminogen activator receptor (suPAR) as an early predictor of severe respiratory failure in patients with COVID-19 pneumonia. Crit Care 2020;24(1):187. doi: 10.1186/s13054-020-02897-4

4. Rasmussen U, Ladelund S, Haupt TH, et al. Soluble urokinase plasminogen activator receptor (suPAR) in acute care: a strong marker of disease presence and severity, readmission and mortality. A retrospective cohort study. Emerg Med J 2016;33(11):769–75. doi: 10.1136/emermed-2015-205444

5. Rasmussen UH, Ladelund S, Haupt TH, et al. Combining National Early Warning Score With Soluble Urokinase Plasminogen Activator Receptor (suPAR) Improves Risk Prediction in Acute Medical Patients: A Registry-Based Cohort Study. Crit Core Med 2018;46(12):1961–68. doi: 10.1097/CCM.0000000000003441

6. Hoenigl M, Moser CB, Funderburg N, et al. Soluble Urokinase Plasminogen Activator Receptor Is Predictive of Non-AIDS Events During Antiretroviral Therapy-mediated Viral Suppression. Clin Infect Dis 2019;69(4):676–86. doi: 10.1093/cid/ciy966

7. Oliveira I, Andersen A, Furtado A, et al. Assessment of simple risk markers for early mortality among HIV-infected patients in Guinea-Bissau: a cohort study. BMJ Open 2012;2(6) doi: 10.1136/bmjopen-2012-001587

8. Outinen TK, Tervo L, Makela S, et al. Plasma levels of soluble urokinase-type plasminogen activator receptor associate with the clinical severity of acute Puumala hantavirus infection. PLoS One 2013;8(8):e71335. doi: 10.1371/journal.pone.0071335

9. Yilmaz G, Mentese A, Kaya S, et al. The diagnostic and prognostic significance of soluble urokinase plasminogen activator receptor in Crimean-Congo hemorrhagic fever. J Clin Virol 2011;50(3):209–11. doi: 10.1016/j.jcv.2010.11.014

10. Citlenbik H, Ulusoy E, Er A, et al. Levels of Soluble Urokinase Plasminogen Activator Receptor in Pediatric Lower Respiratory Tract Infections. Pediatr Allergy Immunol Pulmonol 2019;32(3):121–27. doi: 10.1089/ped.2018.0982

11. Gumus A, Altintas N, Cinarka H, et al. Soluble urokinase-type plasminogen activator receptor is a novel biomarker predicting acute exacerbation in COPD. Int J Chron Obstruct Pulmon Dis 2015;10:357–65. doi: 10.2147/COPD.S77654

12. Chen D, Wu X, Yang J, et al. Serum plasminogen activator urokinase receptor predicts elevated risk of acute respiratory distress syndrome in patients with sepsis and is positively associated with disease severity, inflammation and mortality. Exp Ther Med 2019;18(4):2984–92. doi: 10.3892/etm.2019.7931

13. Yang X, Yu Y, Xu J, et al. Clinical course and outcomes of critically ill patients with SARS-CoV-2 pneumonia in Wuhan, China: a single-centered, retrospective, observational study. Lancet Respir Med 2020;8(5):475–81. doi: 10.1016/S2213-2600(20)30079-5

14. Harris PA, Taylor R, Thielke R, et al. Research electronic data capture (REDCap)--a metadata-driven methodology and workflow process for providing translational research informatics support. J Biomed Inform 2009;42(2):377–81. doi: 10.1016/j.jbi.2008.08.010

15. Bachelet VC. Do we know the diagnostic properties of the tests used in COVID-19? A rapid review of recently published literature. Medwave 2020;20(3):e7890. doi: 10.5867/medwave.2020.03.7891

16. Miyara M, Tubach F, Pourcher V, et al. Low incidence of daily active tobacco smoking in patients with symptomatic COVID-19.

17. Schultz M, Rasmussen LJH, Hoi-Hansen T, et al. Early Discharge from the Emergency Department Based on Soluble Urokinase Plasminogen Activator Receptor (suPAR) Levels: A TRIAGE III Substudy. Dis Markers 2019;2019:3403549. doi: 10.1155/2019/3403549

18. Velissaris D, Dimopoulos G, Parissis J, et al. Prognostic Role of Soluble Urokinase Plasminogen Activator Receptor at the Emergency Department: A Position Paper by the Hellenic Sepsis Study Group. Infect Dis Ther 2020:1–10. doi: 10.1007/s40121-020-00301-w

19. Hayek SS, Leaf DE, Samman Tahhan A, et al. Soluble Urokinase Receptor and Acute Kidney Injury. N Engl J Med 2020;382(5):416–26. doi: 10.1056/NEJMoa1911481

20. Iversen E, Houlind MB, Kallemose T, et al. Elevated suPAR Is an Independent Risk Marker for Incident Kidney Disease in Acute Medical Patients. Front Cell Dev Biol 8 2020;8 doi: 10.3389/fcell.2020.00339

21. Eugen-Olsen J, Andersen O, Linneberg A, et al. Circulating soluble urokinase plasminogen activator receptor predicts cancer, cardiovascular disease, diabetes and mortality in the general population. J Intern Med 2010;268(3):296–308. doi: 10.1111/j.1365-2796.2010.02252.x

22. Ji HL, Zhao R, Matalon S, et al. Elevated Plasmin(ogen) as a Common Risk Factor for COVID-19 Susceptibility. Physiol Rev 2020;100(3):1065–75. doi: 10.1152/physrev.00013.2020

23. Christopher M, Petrilli CM, Jones SA, Yang j et al, Factors associated with hospital admission and critical illness among 5279 people with coronavirus disease 2019 in New York City: prospective cohort study. BMJ 2020;369:m1966

